# Megakaryocytes are a Novel SARS-CoV-2 Infection Target and Risk Factor for Mortality and Multi-Organ Failure

**DOI:** 10.1101/2021.08.05.21261552

**Authors:** Seth D. Fortmann, Michael J. Patton, Blake F. Frey, Cristiano P. Vieira, Sivani B. Reddy, Forest Huls, Andrew Crouse, Jason Floyd, Ram Prasad, Vidya Sagar Hanumanthu, Sarah Sterrett, Jeremy D. Zucker, Peng Li, Nathan Erdmann, Paul A. Goepfert, Amit Gaggar, Maria B. Grant, Matthew Might

## Abstract

Discovery of a biomarker for patients at high risk of progression to severe Coronavirus Disease 2019 (COVID-19) is critical for clinical management, particularly in areas of the world where widespread vaccine distribution and herd immunity have yet to be achieved. Herein, we characterize peripheral blood from 218 COVID-19 patients with flow cytometry and provide evidence that megakaryocytes are a target for infection by Severe Acute Respiratory Syndrome Coronavirus 2 (SARS-CoV-2). We demonstrate a positive correlation between infected megakaryocytes expressing the protein calprotectin (also called S100A8/A9), a known marker of COVID-19 severity. Additionally, we show that infected, calprotectin expressing megakaryocytes are correlated with COVID-19 severity and are a prognostic indicator of 30-day clinical outcomes including respiratory failure, thrombotic events, acute kidney injury (AKI), ICU admission, and mechanical ventilation. These findings represent a novel SARS-CoV-2 infection target with significant clinical implications as a biomarker for clinical outcomes associated with severe COVID-19.

## Main Text

Severe COVID-19 is a characterized by dysfunction of platelet precursor cells called megakaryocytes. Autopsy reports have demonstrated megakaryocytes in the hearts, kidneys, liver, lungs, and brains of COVID-19 patients, suggesting increased mobilization of megakaryocytes into the peripheral circulation^1–5^. Recent multi-omic experiments performed on peripheral blood from COVID-19 patients have shown elevated levels of circulating megakaryocytes expressing uniquely high levels of calprotectin (S100A8/A9), an antimicrobial protein characteristically expressed by neutrophils and classical monocytes^6–9^. While the presence of calprotectin in immune cell populations has been reported as a marker of COVID-19 severity and has been associated with post-infection cytokine storm^7,10^, little is known about the phenotypes of circulating megakaryocytes in COVID-19 and whether these cells have prognostic value.

To further investigate megakaryocytes as a biomarker of COVID-19 severity, peripheral blood was collected from 218 COVID-19+ patients admitted to University of Alabama at Birmingham (UAB) hospital. The baseline characteristics of the patient cohort are shown in Table 1 (see Table S1 for extended cohort details). The mean age (*±* standard deviation; SD) was 61 *±* 13 years, 58% were male, mean body mass index (BMI) was 34 *±* 10, mean Charlson comorbidity score was 2.8 *±* 2.1, and the average time to sample collection, relative to admission, was 5.9 *±* 3.5 days. Patient COVID-19 disease severity was computed on the day of sample collection using the World Health Organization (WHO) 8-point ordinal scale^11^(Table 1). Using flow cytometry, we isolated circulating megakaryocytes from peripheral blood that were live singlets, positive for megakaryocyte markers CD61 and CD41 (Figure 1A left panel; see Figure S5 for full gating strategy)^12^. Megakaryocytes were distinguished from platelets using propidium iodide DNA staining and CD45 (Figure 1A, middle panel) which revealed ploidy ranging from 2N to 32N (Figure 1A, right panel). Using fluorescent activated cell sorting (FACS), we isolated CD61+, CD41+, DNA+ cells from COVID-19 positive peripheral blood and observed a heterogeneous population of circulating megakaryocytes ranging in size from 20-50 micrometers (Figure 1B). In agreement with reports of peripheral mobilization of megakaryocytes in COVID-19 patients, we found that the COVID-19 positive cohort had a significantly higher volume of circulating megakaryocytes as a percent of all living cells isolated from peripheral blood than uninfected controls (Figure 1C; 218 distinct patients, 2 with multiple sample collection time-points)^1–5^. Using CD45 and propidium iodide staining, we found that circulating megakaryocytes were effectively absent in uninfected control samples (Figure 1D, right panel) compared to COVID-19 infected patients (1D, right panel). Circulating megakaryocytes from COVID-19 positive patients were then stained for calprotectin and SARS-CoV-2 spike protein, a canonical marker of COVID-19 infection. We identified three distinct circulating megakaryocyte populations in COVID-19 infected patients (Figure 1E): double negative circulating megakaryocytes (calprotectin-spike-, Figure 1D, lower-left quadrant), calprotectin positive circulating megakaryocytes (calprotectin+ spike-, Figure 1D, upper-left quadrant), and double positive circulating megakaryocytes (calprotectin+ spike+, Figure 1D, upper-right quadrant)^7^. Using the PrimeFlow assay, we confirmed the simultaneous presence of calprotectin mRNA and protein in the calprotectin+ spike+ circulating megakaryocytes (Figure 1F), providing evidence that megakaryocytes are actively producing antimicrobial proteins in the context of SARS-CoV-2 infection. We then hypothesized that SARS-CoV-2 could be replicating in megakaryocytes and potentially inducing aberrant expression of calprotectin. Flow cytometry imaging of circulating megakaryocyte populations revealed cells with large nuclei characteristic of megakaryocytes that displayed CD61, CD41, calprotectin, and spike protein (Figure 1G^12^. Spike protein staining in the calprotectin+ circulating megakaryocytes exhibited a punctate perinuclear pattern (Figure 1G, red), consistent with SARS-CoV-2 replication in the Golgi complex^13^. In infected circulating megakaryocytes, we observed the formation of possible proplatelets that contained spike protein (Figure 1G, inset white arrow), raising the intriguing possibility that infected megakaryocytes may be a source of SARS-CoV-2 containing platelets. While further studies are required to show passage of virus from megakaryocyte to platelet, our results provide a plausible explanation for the recent paradoxical observation of human platelets containing SARS-CoV-2 RNA without expressing angiotensin-converting enzyme 2 (ACE2) required for viral entry^14^.

**Table 1.**
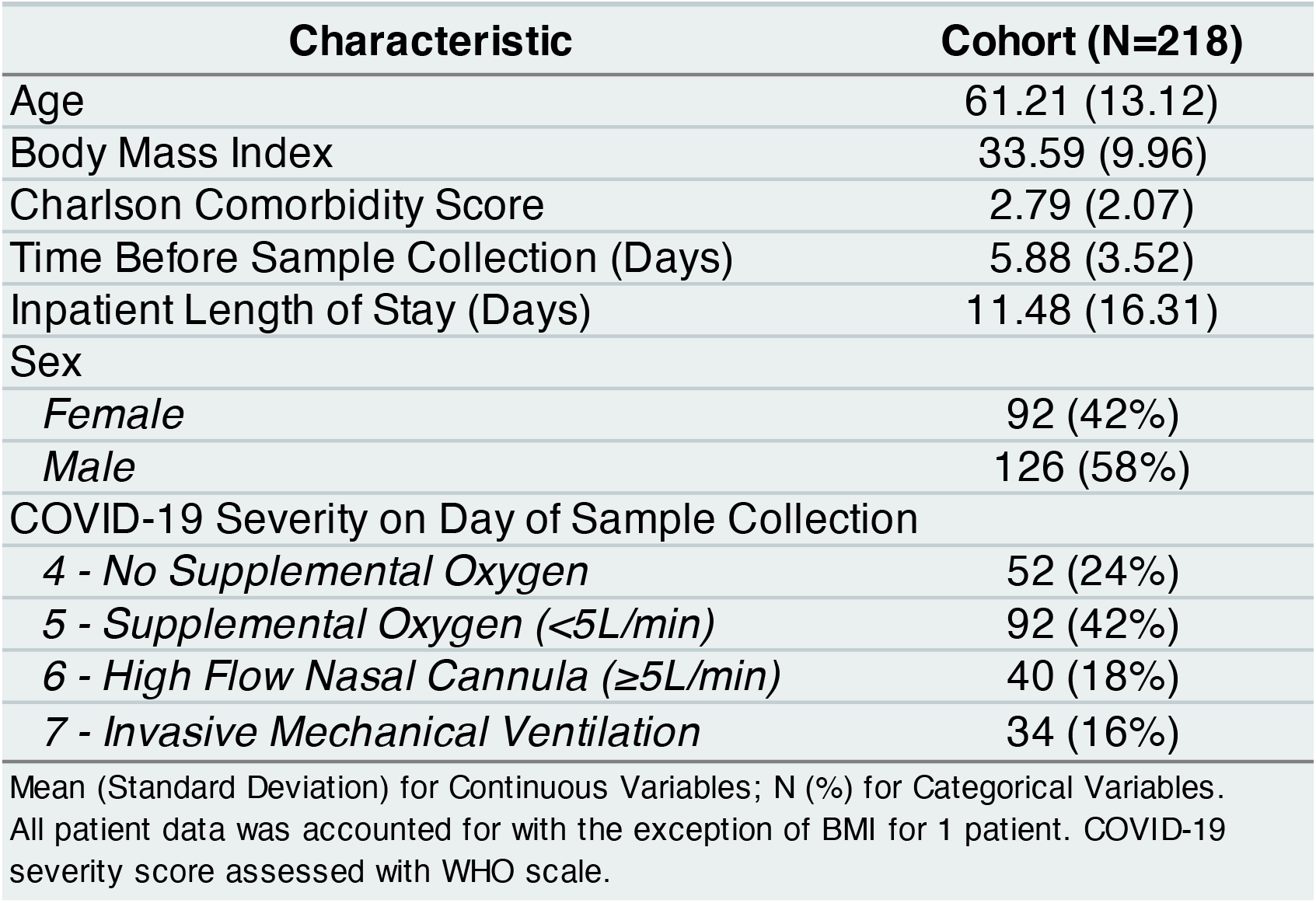
Study Population

**Figure 1:**
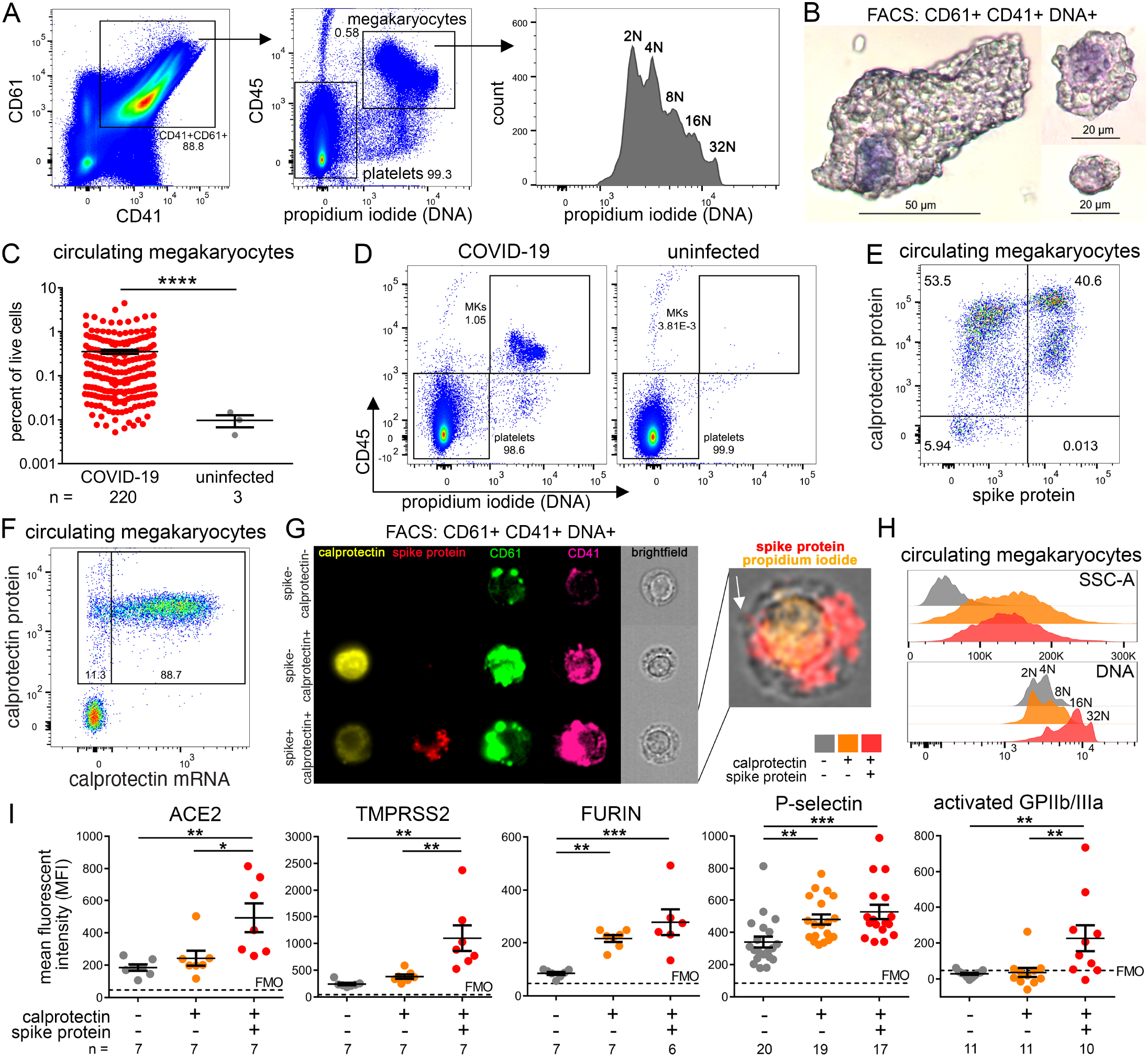
Calprotectin+ Spike+ Megakaryocytes are Infected with SARS-CoV-2. **(A)** Flow cytometry from UAB COVID-19 peripheral blood samples (n=20) showing gating of CD61+, CD41+, CD45+, DNA+ megakaryocytes. DNA- cells in bottom left corner of center panel are platelets. Right panel shows distinct ploidy of megakaryocytes ranging from 2-32N. **(B)** FACS sorted circulating megakaryocytes (CD61+ CD41+ DNA+) from COVID-19 peripheral blood mounted using cytocentrifuge and stained with hematoxylin and eosin. Representative images showing the size/morphology spectrum of circulating megakaryocytes (circulating megakaryocytes). **(C)** Quantification of circulating megakaryocytes (CD61+ CD41+ DNA+) using flow cytometry from COVID-19 (n=220 patients) versus uninfected controls (n=3 donors). Unpaired t test with Welch’s correction. P-value: **** < 0.0001. **(D)** Circulating megakaryocytes from COVID-19+ peripheral blood (n=1) vs. uninfected control blood (n=3) stained for two intracellular protein targets for CD45 and propidium iodide. **(E)** Circulating megakaryocytes from COVID-19+ peripheral blood (n=1) calprotectin (calprotectin; y-axis) and SARS-CoV-2 Spike protein (x-axis). Quadrant gating shows 3 distinct megakaryocyte subpopulations in the COVID-19 positive sample: calprotectin-spike-(lower-left), calprotectin+ spike-(upper-left), and calprotectin+ spike+ (upper-right). **(F)** Representative PrimeFlow flow cytometry plot of circulating megakaryocytes (CD61+ CD41+ DNA+) from a COVID-19 patient (n=1) showing S100A8/A9 protein expression and S100A8/A9 mRNA expression. Target probes to S100A8 and S100A9 were labeled with the same fluorophore. **(G)** Imaging flow cytometry of the three megakaryocyte subpopulations showing images from five channels: S100A8/A9 (yellow), spike protein (red), CD61 (green), CD41 (magenta), and brightfield (grey). Inset shows overlay of brightfield (grey), spike protein (red), and propidium iodide (DNA; orange) from the S100A8/A9+ spike protein+ megakaryocytes; white arrow designates possible proplatelet formation. **(H)** Side scatter area (SSC-A; granularity) and propidium iodide (DNA) histogram plots of the three circulating megakaryocyte subpopulations found in COVID-19+ patients (n=20). **(I)** Mean fluorescent intensity (MFI) analyses. For MFI of ACE2, TM-PRSS2, and FURIN, all three antibodies were conjugated to Alexa Fluor 488, and the same samples (n=7 patients) were stained separately with the three antibodies. For P-selectin analysis, n=20 patients. For P-selectin and GPIIb/IIIa, some COVID-19 patients lacked all 3 megakaryocyte subpopulations. MFI statistical analyses were done using one-way ANOVA with Tukey’s post-hoc multiple comparisons test; adjusted p-values: * = 0.05-0.01, ** = 0.099-0.001, *** = <0.001.

We next sought to characterize the phenotypes of infected circulating megakaryocytes in COVID-19. Calprotectin+ spike- and calprotectin+ spike+ circulating megakaryocytes had increased side scatter values, suggesting increased cell granularity and consistent with calprotectin packaging in granules (Figure 1H, top panel, orange and red)^15^. Interestingly, the calprotectin+ spike+ circulating megakaryocyte population showed right-shifted, multi-peaked propidium ioidide signal consistent with 16-32N chromosomal copies (Figure 1H, bottom panel, red) compared to calprotectin+ spike- and calprotectin-spike-populations which were predominantly 2-8N (Figure 1H, bottom panel, orange and grey). Whether SARS-CoV-2 preferentially infects polyploid megakaryocytes or SARS-CoV-2 promotes DNA replication in infected megakaryocytes remains to be determined. To determine the susceptibility of megakaryocytes to SARS-CoV-2 infection, we investigated three proteins involved in SARS-CoV-2 infection of host cells: 1) ACE2, 2) transmembrane protease serine 2 (TM-PRSS2), 3) FURIN protease^13^. We found that ACE2 and TMPRSS2 had increased expression in the calprotectin+ spike+ megakaryocytes (Figure 1I). FURIN was significantly upregulated in both the calprotectin+ spike+ and calprotectin-spike+ circulating megakaryocytes (Figure 1I). In addition to viral receptors, we also stained for two markers of megakaryocyte activation (P-selectin and activated GPIIb/IIIa), both of which had increased expression in the calprotectin+ spike+ circulating megakaryocyte population (Figure 1I). Taken in sum, these findings represent the strongest evidence to date for megakaryocytes as a novel SARS-CoV-2 infection target.

To evaluate the clinical significance of circulating megakaryocytes, we next examined COVID-19+ acute respiratory distress syndrome (ARDS) lung specimens. We observed DNA+ (Hoescht stain), CD61+, calprotectin+, spike+ megakaryocytes in COVID-19 ARDS lung tissue (Figure 2A), confirming the presence of infected calprotectin+ megakaryocytes in the parenchyma of diseased organs in addition to peripheral circulation. We then investigated the relationship between calprotectin+ spike+ circulating megakaryocyte proportion, laboratory measurements, and adverse clinical outcomes. Spearman correlation analyses were performed to compare calprotectin+ spike+ circulating megakaryocyte proportion to complete blood count (CBC) measurements made within 48 hours of blood sample collection (Figure 2B). We found that calprotectin+ spike+ circulating megakaryocyte proportion, monocyte count (R=-0.35, p-value<0.001, Figure 2B), and lymphocyte count (R=-0.54, p-value<0.001, Figure 2B) were negatively correlated. Calprotectin+ spike+ circulating megakaryocyte proportion was positively correlated with neutrophil count (R=0.52, p-value<0.001, Figure 2B). Lymphopenia and neutrophilia are widely recognized CBC measurements indicative of severe COVID-19 infection, suggesting that calprotectin+ spike+ circulating megakaryocyte may be an additional biomarker of COVID-19 severity^16^. In agreement, we observed a significantly greater calprotectin+ spike+ megakaryocyte proportion in patients with higher WHO COVID-19 severity scores compared to those on room air at the time of sample collection (Figure 2C, red) and at each patient’s worst severity score recorded during their entire inpatient stay (Figure 2C, blue).

**Figure 2:**
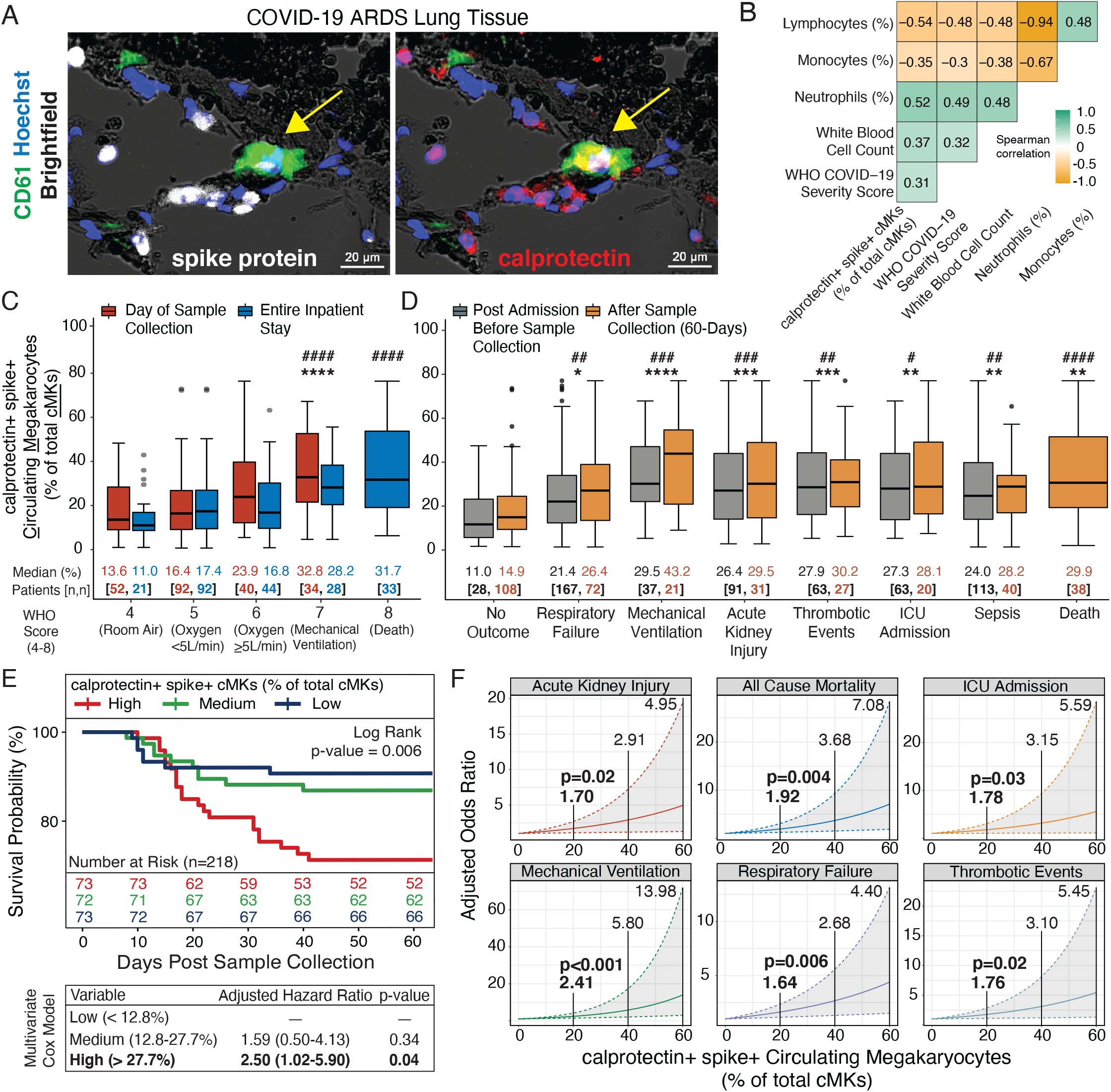
Infected Calprotectin+ Megakaryocytes are an Independent Risk Factor for COVID-19 Mortality and Multi-Organ Injury. **(A)** Immunofluorescence staining of lung tissue from a deceased COVID-19 patient with acute respiratory distress syndrome (ARDS). 5 channels are shown: brightfield (black pigment from TrueView autofluorescence quencher), green (CD61), red (calprotectin), white (spike protein), and blue (Hoechst). Yellow arrows indicates a CD61+ calprotectin+ spike+ megakaryocyte; arrow tail points to lumen of a large vessel. **(B)** Spearman correlation analysis comparing calprotectin+ spike+ circulating megakaryocyte (cMK) proportion (% of total cMKs) to COVID-19 severity (WHO 8 point ordinal scale) and immune cell frequencies assessed within 48 hours of sample collection. Patients with multiple laboratory values during that time frame were averaged. **(C)** Analyses comparing COVID-19 severity and calprotectin+ spike+ cMK proportion. COVID-19 severity on the day of sample collection (red) and the worst severity score during the entire inpatient stay (blue). Median values for calprotectin+ spike+ cMK proportion and color coded patient counts are displayed below each distribution. **(D)** Analyses comparing COVID-19 adverse events and calprotectin+ spike+ cMK proportion prior to sample collection (grey) and 60-days after sample collection (yellow). Statistical analyses were done comparing pre-sample collection outcomes versus no pre-sample collection outcomes (*) and 60-day post-sample collection outcomes versus no 60-day outcomes (#). Median values and color-coded patient counts are reported below each distribution. Statistical significance for (C-D) was assessed with the Dunn test and adjusted for multiple comparisons with the Bonferroni method (adjusted p-value: */# = 0.05-0.01, **/## = 0.01-0.001, ***/### = 0.001-0.0001, ****/#### = <0.0001). **(E)** 60-day Kaplan-Meier survival curves subset by calprotectin+ spike+ circulating MK proportions (equivalent patient numbers per distribution: high ≥27.7%, medium 12.8-27.7%, low <12.8%) with accompanying adjusted hazard ratio table derived from multivariate Cox regression model. Survival curve statistical significance determined using log-rank test comparing high group to medium and low groups. **(F)** Multivariate logistic regression models with calprotectin+ spike+ cMK proportion as a continuous variable showing likelihood of 30-day outcomes (acute kidney injury, all-cause mortality, ICU admission, mechanical ventilation, respiratory failure, and thrombotic events). Confidence intervals denoted by grey shaded region in each plot, with adjusted odds ratios estimates annotated above the 20% (p-value included), 40%, and 60% intervals. Sepsis was not statistically significant and therefore, is not shown graphically. Statistical significance for the Cox regression model (E) and multivariate logistic regression models (F) was assessed using the companion applied regression analysis-of-variance (CAR-ANOVA) type III test.

We next investigated specific adverse outcomes at two distinct time intervals during each patient’s COVID-19 inpatient stay: 1) post-admission and prior to sample collection and 2) within 60 days following sample collection (see detailed outcome distribution in Table S2, Figure S1, S2, S3, S4)). Outcomes prior to sample collection (Figure 2D, grey) showed a statistically significant (median, adjusted p-value = *) increase in calprotectin+ spike+ circulating megakaryocyte proportion for respiratory failure (21.4%, p=0.03), mechanical ventilation (29.5%, p<0.0001), acute kidney injury (AKI; 26.4%, p=0.001), thrombotic events (27.9%, p=0.001), intensive care unit (ICU) admission (27.3%, p=0.002), and sepsis (24.0%, p=0.004) compared to those without pre-sample collection outcomes (11.0%; Figure 2D). Importantly, statistically significant elevations in calprotectin+ spike+ circulating megakaryocyte proportion were also found in patients with 60-day post sample collection outcomes (2D, gold, adjusted p-value = #): acute respiratory failure (26.4%, p=0.003), mechanical ventilation (43.2%, p=0.001), thrombotic events (30.2%, p=0.004), AKI (29.5%, p=0.008), ICU admission (28.1%, p=0.026), sepsis (28.2%, p=0.008), and death (29.9%, p<0.0001) compared to those without a 60-day outcome (14.9%; Figure 2D).

We then categorized patients into three equally distributed groups based on the cohort distribution of calprotectin+ spike+ circulating megakaryocyte proportions (high *≥* 27.7%, medium 12.8-27.7%, low <12.8%) and performed a 60-day survival analysis using the Kaplan-Meier method. Patients in the high calprotectin+ spike+ circulating megakaryocyte group experienced the greatest reduction in 60-day survival probability (log-rank test p=0.006; Figure 2E; see Figure S14 for complete survival timeline). Multivariate Cox regression analysis controlling for age, BMI, time to sample collection, comorbidity score, and sex, confirmed a statistically significant adjusted hazard ratio (HR) increase for patients in the high group compared to the low group (adjusted HR:2.50, 95% CI:1.02-5.90, p=0.04; Figure 2E, see Figure S3 for model details). Using multivariate logistic regression with calprotectin+ spike+ megakaryocyte proportion as a continuous variable, we observed similar trends when this analysis was extended to a more stringent 30-day outcome window (2F, see Table S4 model details). For every 20% increase in calprotectin+ spike+ circulating megakaryocyte proportion, adjusted odds ratios for each outcome were (adjusted OR, 95% CI, p-value): AKI (1.70, 1.09-2.69, p=0.02), all-cause mortality (1.92, 1.23-3.05, p=0.004), ICU admission (1.78, 1.03-3.06, p=0.03), mechanical ventilation (2.41, 1.43-4.16, p<0.001), respiratory failure (1.64, 1.15-2.36, p=0.006) and thrombotic events (1.76, 1.09-2.85, p=0.02) (see Figure 2F, bold). The independent association of these infected megakaryocytes with metrics of clinical outcomes, ICU stay, and target organ injury suggests a central role for these cells in the pathology of severe SARS-CoV-2 infection.

Using multiple methods, our study establishes megakaryocytes as a novel target of SARS-CoV-2 infection, expressing ACE2, TMPRSS2, and FURIN to facilitate viral entry. Further, these infected megakaryocytes contain uniquely high levels of calprotectin (S100A8/A9), an antimicrobial protein characteristically expressed by neutrophils and classical monocytes, and found to be elevated in myeloid populations in severe COVID-19^10^. Our work not only highlights infected megakaryocytes in circulation but also shows them in target organs, such as the lung. The identification of this cell population in critical illness provides the strong possibility that they may serve as biomarkers in other ICU-related disorders. Two noteworthy shortcomings of our study were a single time-point per patient for blood collection and differing post admission collection times for each patient. The latter shortcoming (time to collection) was controlled for in both the multivariate Cox hazard ratio and logistic regression odds ratio models. Nevertheless, identification of these cells in severe COVID-19 may inform new therapeutic targets to improve outcomes in these patients. To our knowledge, this represents the first biomarker in COVID-19 subjects with associations across multiple metrics of disease severity. Moreover, this work constitutes the first evidence of megakaryocyte infection by any respiratory virus and suggest that megakaryocytes may be susceptible to other viral pathogens in the coronavirus family.

## Data Availability

Upon acceptance of the manuscript for publication, an UAB approved subset of de-identified data pertaining to the analyses within the manuscript will be made available.

## Author Contributions (names appear alphabetized by last name)

**Conceptualization:**

Seth D. Fortmann, Michael J. Patton

**Methodology:**

Seth D. Fortmann, Blake F. Frey, Michael J. Patton

**Investigation:**

Jason Floyd, Seth D. Fortmann, Blake F. Frey, Vidya Sagar Hanumanthu, Michael J. Patton, Sivani B. Reddy, Cristiano P. Vieira

**Institutional Review Board Protocol Design:**

Andrew Crouse, Maria B. Grant, Michael J. Patton

**Data Visualization:**

Seth D. Fortmann, Peng Li, Michael J. Patton

**Statistical Analyses:**

Seth D. Fortmann, Peng Li, Michael J. Patton, Jeremy D. Zucker

**Funding Acquisition:**

Andrew Crouse, Maria B. Grant, Matthew Might

**Original Draft Writing & Compilation:**

Michael J. Patton

**Draft Review & Editing:**

All Co-authors

