## supplemental_materials for "Megakaryocytes are a Novel SARS-CoV-2 Infection Target and Risk Factor for Mortality and Multi-Organ Failure"

Any methods, additional references, reporting summaries, source data, extended data, supplementary information, acknowledgements, peer review information; details of author contributions and competing interests; and statements of data and code availability are available at: https://github.com/TriageDr.

### Methods

#### Study Population and Electronic Medical Record Data Collection

Between August 2020 and March 2021, 1,961 patients who were hospitalized at University of Alabama at Birmingham (UAB) Hospitals with COVID-19 and had a positive Severe Acute Respiratory Syndrome Coronavirus 2 (SARS-CoV-2) PCR test were enrolled in the Enterprise study. Of these 1,961 patients, peripheral blood was collected from 619, and 218 were included in this study. Written informed consent was obtained from all patients, and the study protocol was IRB approved (UAB; IRB-300006291). Patient demographics, past medical history, biometrics, laboratory measurements, and all clinical outcomes were obtained via electronic medical records (EMR) from UAB. Charlson comorbidity indices were constructed for each patient using EMR data collected prior to their COVID-19 encounter. Peak COVID-19 disease severity was assessed using the World Health Organization (WHO) ordinal scale across two time intervals: the day of sample collection and the entire inpatient stay (Table 1S). The following clinical outcomes: all-cause mortality, acute respiratory failure, mechanical ventilation, thrombotic events (venous and arterial), sepsis, acute kidney injury (AKI), and ICU admission, were identified using International Code of Disease 10 (ICD-10) identifiers (Table 2S, and were assessed at two time intervals: after COVID-19 admission and before sample collection, and within 60-days after sample collection (for detailed breakdown of each clinical outcome ICD-10 code at each time-point, see the following figures: Figure 1S, Figure 2S, Figure 3S, Figure 4S). ICU admission and mechanical ventilation were determined by admission and procedure time stamps.

#### Peripheral Blood Collection and Processing

Venous blood was drawn from the median cubital vein in acid citrate dextrose coated tubes under sterile conditions. Platelet-rich plasma was separated by centrifugation at 300xg for 10 minutes. Platelet-rich plasma was further centrifuged at 1,200xg for 10 minutes, forming a cell pellet. The cell pellets were resuspended in 1mL of 90% fetal bovine serum (FBS) with 10% dimethyl sulfoxide and stored at -80C for $\leq$6 months until time of analysis.

#### Flow Cytometry

Cryopreserved cell suspensions were rapidly thawed in a 37C water bath for 3 minutes and filtered through a 100m strainer. 1mL of fresh FBS was added to each sample followed by centrifugation at 1000xg for 5 minutes at 4oC. The samples were resuspended in 1mL of flow cytometry staining buffer (FACS; phosphate-buffered saline with 5% FBS, 2mM EDTA, and 0.1% sodium azide). 10L of each sample was stained with anti-CD61 (VI-PL2; AF647) and the concentration of CD61+ cells were determined using flow cytometry. Ten million CD61+ cells were used to standardize each sample for the final flow cytometry panel. Cell suspensions were surface stained at room temperature for 30 minutes in 100L of staining buffer (50L of Brilliant Stain Buffer (Becton Dickinson) and 50L of FACS) with the following antibody cocktail: anti-CD61 (VI-PL2; BV605), anti-CD41 (HIP8; APC/Cy7), anti-CD45 (HI30; BV510), viability dye (violet; Invitrogen), and Human TruStain FcX (BioLegend). Cells were fixed and permeabilized with BD Cytofix/Cytoperm (Becton Dickinson) for 30 minutes on ice. Cells were intracellularly stained with anti-calprotectin (27E10; Santa Cruz) and anti-spike protein (P05DHuRb; Invitrogen) for 30 minutes on ice in 100L of Perm/Wash Buffer (Becton Dickinson). Finally, cells were incubated with 200L RNase and propidium iodide (PI/RNase Staining Buffer; Becton Dickinson) for 15 minutes at room temperature. Each sample was resuspended in 300L of FACS buffer and analyzed with a three-laser FACSCelesta flow cytometer (Becton Dickinson). All 218 peripheral blood samples were analyzed in 6 batches during a 1 week period. FCS files were analyzed using FlowJo (version 10.7). The gating scheme (Figure S5) and fluorescence minus one (FMO; Figure S6) controls are provided.

For mean fluorescent intensity (MFI) analyses of ACE2, TMPRSS2, and FURIN, the following conjugated antibodies were used: anti-ACE2 (FAB933G[Novus]; AF488), anti-TMPRSS2 (H-4; AF488), and anti-FURIN (B-6; AF488). 7 samples were stained in total, and the same samples were stained three times, once with each of the respective ACE2/TMPRSS2/FURIN antibodies. ACE2 and TMPRSS2 were surface stained while FURIN was intracellularly stained. Statistical analyses of mean MFI values were done using one-way ANOVA with Tukey's post-hoc multiple comparisons test (GraphPad Prism, version 6.01). Grubbs' test (GraphPad Prism) was used to identify one high outlier value in the FURIN group belonging to the double positive megakaryocyte population. Exclusion of this datapoint did not affect statistical significance and was removed from the final analysis to improve plotting. Statistical analyses of mean MFI values were done using one-way ANOVA with Tukey's post-hoc multiple comparisons test (GraphPad Prism, version 6.01).

### Prime Flow

PrimeFlow (ThermoFisher) was performed according to the manufacturer's instructions with minor modifications. Predesigned RNA probes to human S100A8 and S100A9 were purchased from ThermoFisher and both were assigned to the Alexa Fluor 750 channel. Premade RNA probes to the SARS-CoV-2 genome were purchased from ThermoFisher and assigned to the Alexa Fluor 488 (AF488) channel. 30 million CD61+ cells were used for each sample. Cells were surface stained with anti-CD61 (VI-PL2; BV605), anti-CD41 (HIP8; BV785), anti-CD45 (HI30; BV510), viability dye (violet; Invitrogen), and Fc blocker (Human TruStain FcX; BioLegend). Cells were fixed and permeabilized according to manufacturer's instructions and were intracellularly stained with anti-calprotectin (27E10; Santa Cruz) and anti-spike protein (P05DHuRb; Invitrogen). RNA primer hybridization steps were performed at 40 in a thermal cycler in PCR tubes. After the final RNA fluorophore labeling step, the cells were stained with propidium iodide (eBioscience; 00-6990) for 15 minutes at room temperature, without RNase treatment. PrimeFlow samples were analyzed on a three laser FACSCelesta flow cytometer (Becton Dickinson), and laser voltages were lowered to optimize signal. FCS files were analyzed using FlowJo (version 10.7), and the same general gating scheme was used (Figure 5S). PrimeFlow fluorescence minus one (FMO) controls are provided (Figure 8S).

### FACS Sorting and Imaging Flow Cytometry

Residual samples that were stained for the 218 patient cohort were pooled together and used for fluorescence activated cell sorting (FACS) and imaging flow cytometry experiments. The three megakaryocyte subpopulations, double negative megakaryocytes (calprotectin- spike-), single positive megakaryocytes (calprotectin+ spike-), and double positive megakaryocytes (calprotectin+ spike+), were FACS sorted using a FACSAria II (Becton Dickinson). The cells were then pelleted with centrifugation and 3,000 cells per subpopulation were resuspended in 20L of FACS buffer (phosphate buffered saline with 5% FBS, 2mM EDTA, and 0.1% sodium azide). The samples were run on an Amnis ImageStreamX Mk II System and were analyzed using the IDEAS software (version 6.2). Mouse splenocytes were used for compensation controls. Identical image display mapping values (X range: min & max and midpoint: x & y) for each specific channel were uniformly applied to the three megakaryocyte samples before the final images were exported.

### Immunofluorescence Staining of Lung Tissue

For immunofluorescence staining, lungs from a deceased COVID-19 donor with acute respiratory distress syndrome (ARDS) were inflated isobarically with 10% formalin and preserved in paraffin blocks. Sequential tissue sections of 5m were cut. Paraffin-embedded sections were incubated for 2 hours at 60C. Deparaffinization and rehydration of the slides was performed with three sequential 5 minute incubations in xylene, two sequential 5 minute incubations in 100% ethanol, two sequential 5 minute incubations in 95% ethanol, and washed in distilled water in three sequential 5 minute incubations with gentle agitation. Citrate buffer (Vector Laboratories) prewarmed to 70C was used for antigen retrieval and was incubated in a heated steamer for 20 minutes, followed by three washes in distilled water with gentle agitation. Sections were incubated with 1X phosphate buffered saline (PBS) for 10 minutes and were then blocked with Trident Universal Protein Blocking Reagent (GeneTex; GTX30963) and Human TruStain FcX (Biolegend) at room temperature for 45 minutes. Sections were then stained overnight in Trident Universal Protein Blocking Reagent at 4C with the following conjugated antibodies: anti-CD61 (FITC; VI-PL2), anti-calprotectin (PE; 27E10; Santa Cruz), and anti-spike protein (AF647; P05DHuRb; Invitrogen). Sections were washed with gentle agitation at room temperature using excess 1X Tris-Buffered Saline with 0.01% Triton x-100. Nuclear staining was done using Hoechst. Lastly, the sections were treated with TrueVIEW Autofluorescence Quencher (Vector Laboratories) following the manufacturer's instructions and were mounted using ProLong Gold Antifade Mountant (Invitrogen). Images were collected using a Zeiss Axio Imager Z2 upright microscope. Negative control stains were done using a mouse isotype antibody conjugated to AF488 (Invitrogen; Figure 9S).

### Statistical Analyses of Clinical Correlations and Outcomes

For analyses comparing calprotectin+ spike+ megakaryocyte proportions with COVID-19 severity and clinical outcomes, statistical significance was assessed using the Dunn test with median values from each nonparametric distribution and adjusted for multiple comparisons with the Bonferroni method (see Figure 10S, Figure 11S, Figure 12S, Figure 13S for complete statistical analysis details). Associations between the proportion of calprotectin+ spike+ megakaryocytes and patient survival were determined using the Kaplan–Meier method. Patients were evenly distributed into three categories based on their calprotectin+ spike+ megakaryocyte proportion (High  27.7%, Medium 12.8-27.7%, Low <12.8%). The log-rank test was used to determine statistically significant differences in 60-day survival between the three groups (see Figure 14S for complete survival analysis). Multivariate proportional-hazards regression analysis was performed using the Cox model to determine the association between calprotectin+ spike+ megakaryocyte proportion and 60-day survival. Analyses were adjusted for age, sex, Charlson comorbidity score, body mass index (BMI), and time to sample collection. Adjusted hazard ratios represent the predicted change in hazard from Low to Medium to High (see Figure 3S for model details). Univariate and multivariate logistic regression models were used to determine the association between calprotectin+ spike+

megakaryocyte proportions and the likelihood of 30-day clinical outcomes. Unadjusted and adjusted odds ratios represent the predicted change in likelihood for a 20% increase in calprotectin+ spike+ megakaryocytes (see Figure 4S for model details). Statistical significance for the Cox regression and multivariate logistic regression was assessed using the companion applied regression analysis-of-variance (CAR-ANOVA) type III test. All figures, tables, modeling, and correlation analyses were performed and generated using R version 4.04. Corresponding code will be openly available online at time of publication.

### Supplemental Figures

| Complete Study Population Characteristics | |
|---|---|
| **Characteristic** | **Cohort (N=218)** |
| Age | 61.21 (13.12) |
| Body Mass Index | 33.59 (9.96) |
| Charlson Comorbidity Score | 2.79 (2.07) |
| Time Before Sample Collection (Days) | 5.88 (3.52) |
| Inpatient Length of Stay (Days) | 11.48 (16.31) |
| Sex | |
| *Female* | 92 (42%) |
| *Male* | 126 (58%) |
| COVID-19 Severity on Day of Blood Sample Collection | |
| *4 - No Supplemental Oxygen* | 52 (24%) |
| *5 - Supplemental Oxygen (<5L/min)* | 92 (42%) |
| *6 - High Flow Nasal Cannula (≥5L/min)* | 40 (18%) |
| *7 - Invasive Mechanical Ventilation* | 34 (16%) |
| Race | |
| *American Indian or Alaska Native* | 1 (0.5%) |
| *Asian* | 5 (2.3%) |
| *Black or African American* | 79 (36%) |
| *Hispanic or Latino* | 4 (1.8%) |
| *Multiple* | 1 (0.5%) |
| *White* | 123 (56%) |
| *Decline/Refuse* | 4 (1.8%) |
| *Other* | 1 (0.5%) |
| Medical History Prior to COVID-19 Admission | |
| *Myocardial Infarction* | 69 (32%) |
| *Congestive Heart Failure* | 71 (33%) |
| *Peripheral Vascular Disease* | 52 (24%) |
| *Cerebral Vascular Disease* | 39 (18%) |
| *Chronic Pulmonary Disease* | 71 (33%) |
| *Diabetes without Complications* | 75 (34%) |
| *Diabetes with Complications* | 53 (24%) |
| *Renal Disease* | 60 (28%) |
| *Cancer* | 25 (11%) |
| *Liver Disease* | 8 (3.7%) |
| *HIV/AIDS* | 5 (2.3%) |

Mean (Standard Deviation) for Continuous Variables; N (%) for Categorical Variables. All patient data was accounted for with the exception of Body Mass Indexe (BMI) for 1 patient. COVID-19 severity score assessed with WHO scale. Charlson comorbidity scores were calculatd based on medical history prior to each patient's COVID-19 admission.

**Table 1: Complete Study Population Characteristics.**

| Clinical Outcomes | ICD10 Codes |
|---|---|
| Thrombotic Events | I21.A1, I21.4, I63.9, I51.3, I24.0, I26.99, I82.409, I26.94, I82.401, I82.629, I82.413, I82.412, I21.01, I21.19, I22.2, I82.90, I82.411, I26.93, I82.403, I82.432, I82.811, I63.511, I21.3, I82.442, I82.452, I82.C11, I82.A13, I82.B12, I82.612, T82.868A, G45.9, I63.40, I82.621, I82.441, I82.431, I82.451, I82.C12, I82.210, D73.5, I81, I82.402, K76.3, N28.0, I63.431, I63.113, I21.9, I63.541, I21.11, I82.B11, I82.813, I82.A11, I82.611 |
| Acute Kidney Injury | N17.9, N19, N17.0 |
| Sepsis | A41.89, A41.9, R65.20, R65.21, R78.81, A41.59, B37.7, A41.50, A41.81, A41.02, A40.8, A41.1, A41.52, A41.51, A41.53 |
| Respiratory Failure | J96.01, J80, J96.21, J96.91, J96.90, J96.00, R06.03, J96.02, J96.92, J96.22 |

**Table 2: ICD10 Codes Used for Clinical Outcomes.**

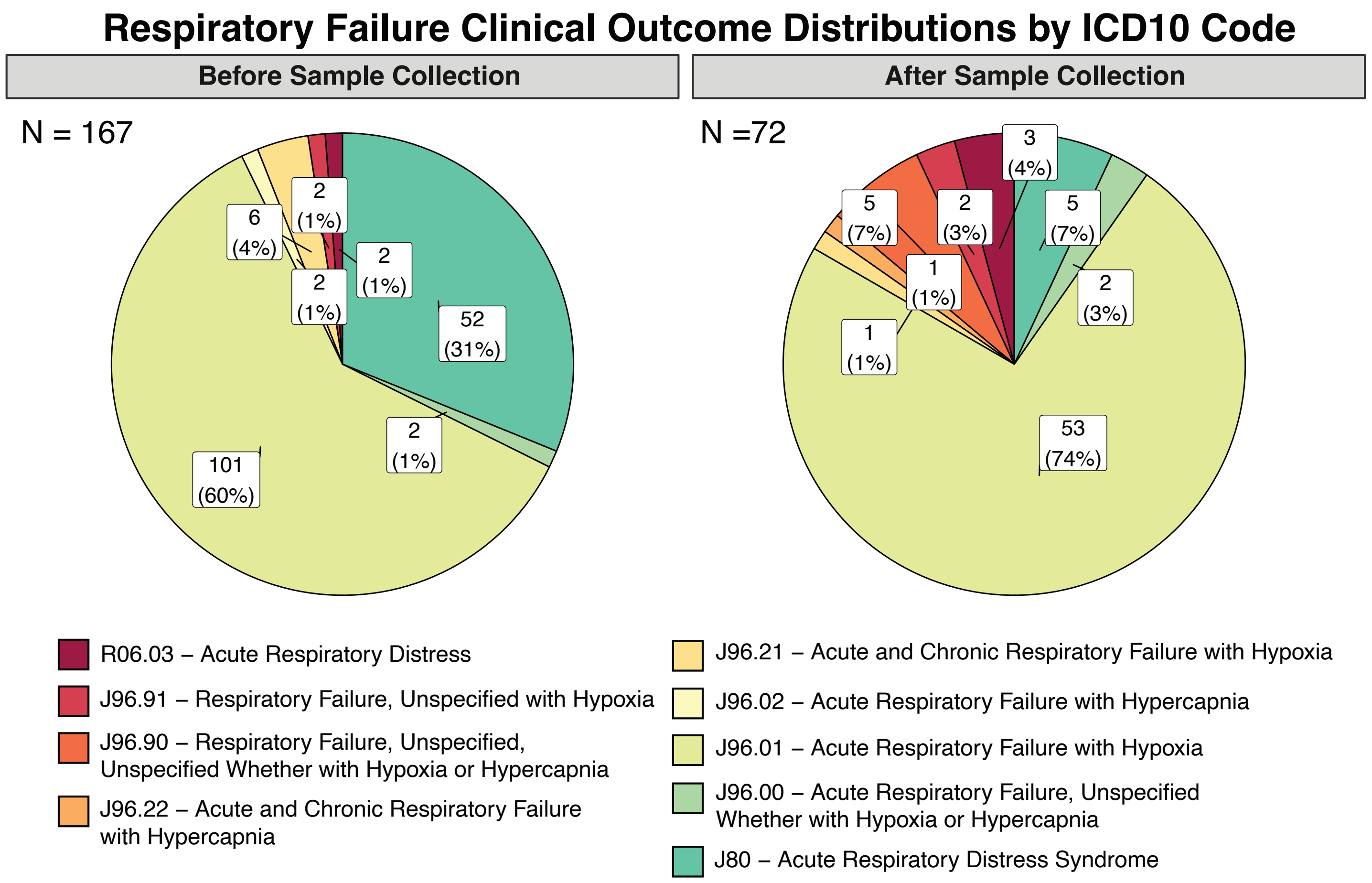


**Figure 1: Distribution of Respiratory Failure Clinical Outcomes**. ICD-10 code derived outcomes were assessed at two time-points: 1) the first respiratory failure outcome found post COVID-19+ admission and prior to blood sample collection, 2) the first respiratory failure outcome found post COVID-19+ admission, within 60-days after blood sample collection. Total outcome count for each time-point (N) are noted in the upper left hand corner of each pie-chart.

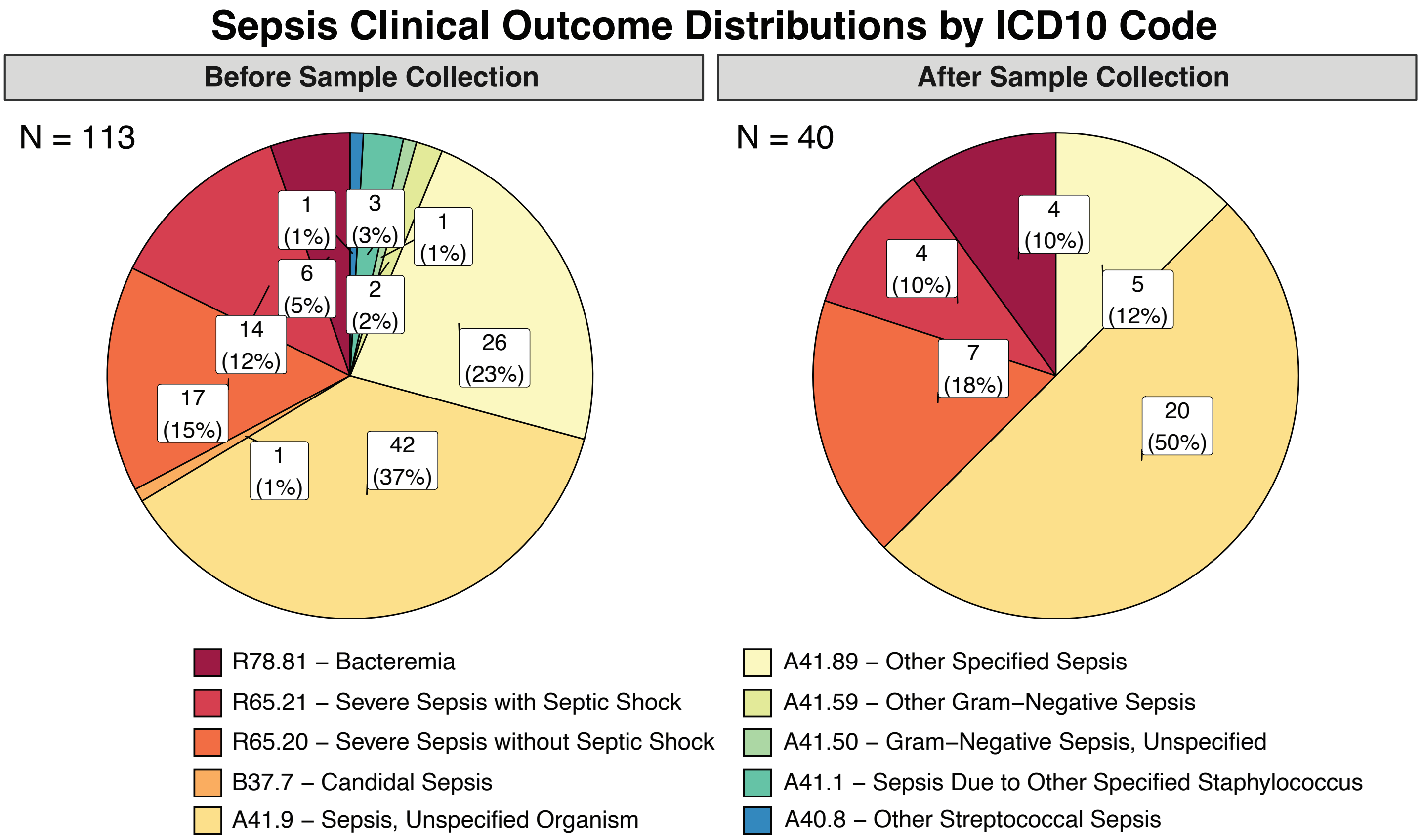


**Figure 2: Distribution of Sepsis Clinical Outcomes**. ICD-10 code derived outcomes were assessed at two time-points: 1) the first sepsis outcome found post COVID-19+ admission and prior to blood sample collection, 2) the first sepsis outcome found post COVID-19+ admission, within 60-days after blood sample collection. Total outcome count for each time-point (N) are noted in the upper left hand corner of each pie-chart.

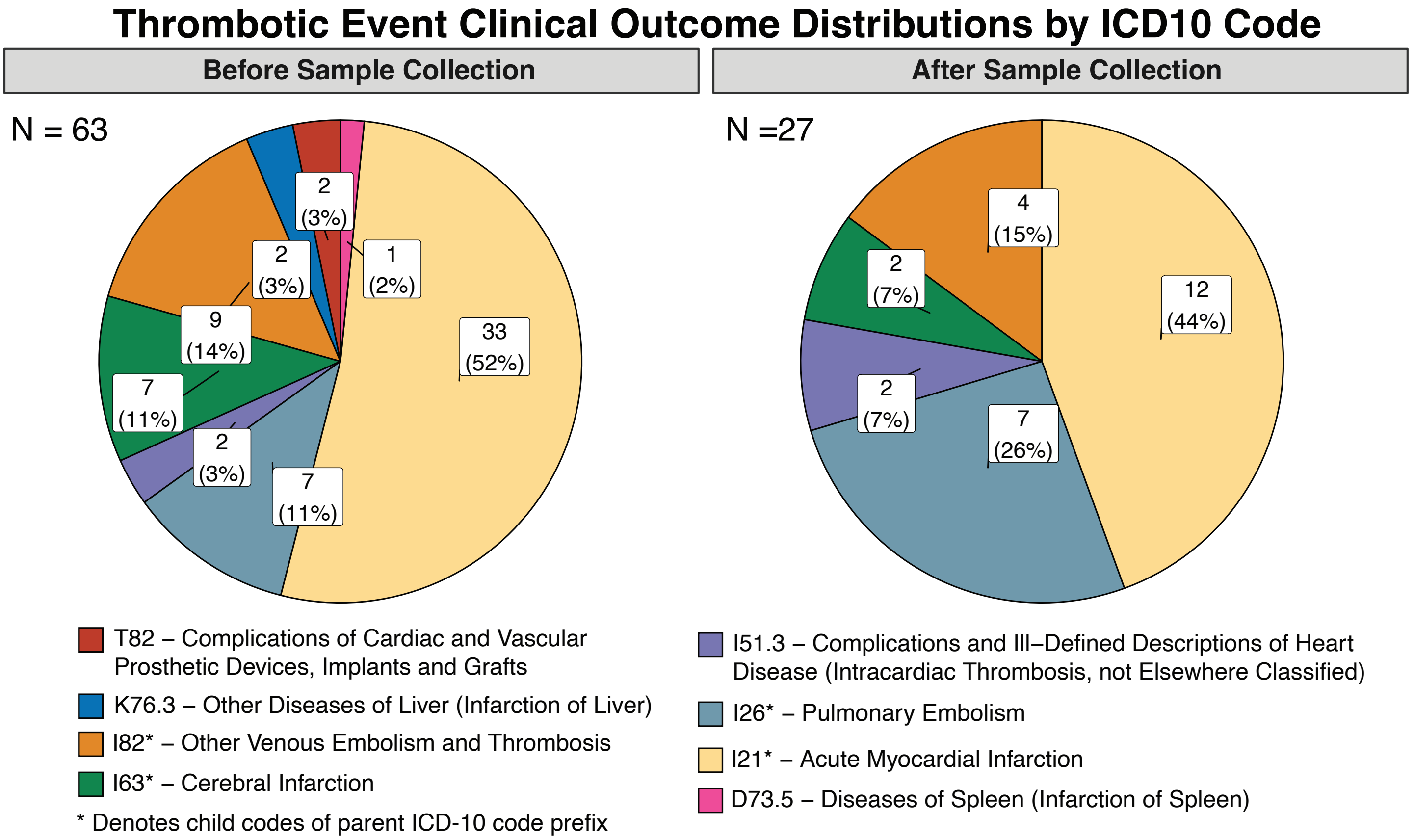


**Figure 3: Distribution of Thrombotic Event Clinical Outcomes**. ICD-10 code derived outcomes were assessed at two time-points: 1) the first thrombotic event outcome found post COVID-19+ admission and prior to blood sample collection, 2) the first thrombotic event outcome found post COVID-19+ admission, within 60-days after blood sample collection. Total outcome count for each time-point (N) are noted in the upper left hand corner of each pie-chart.

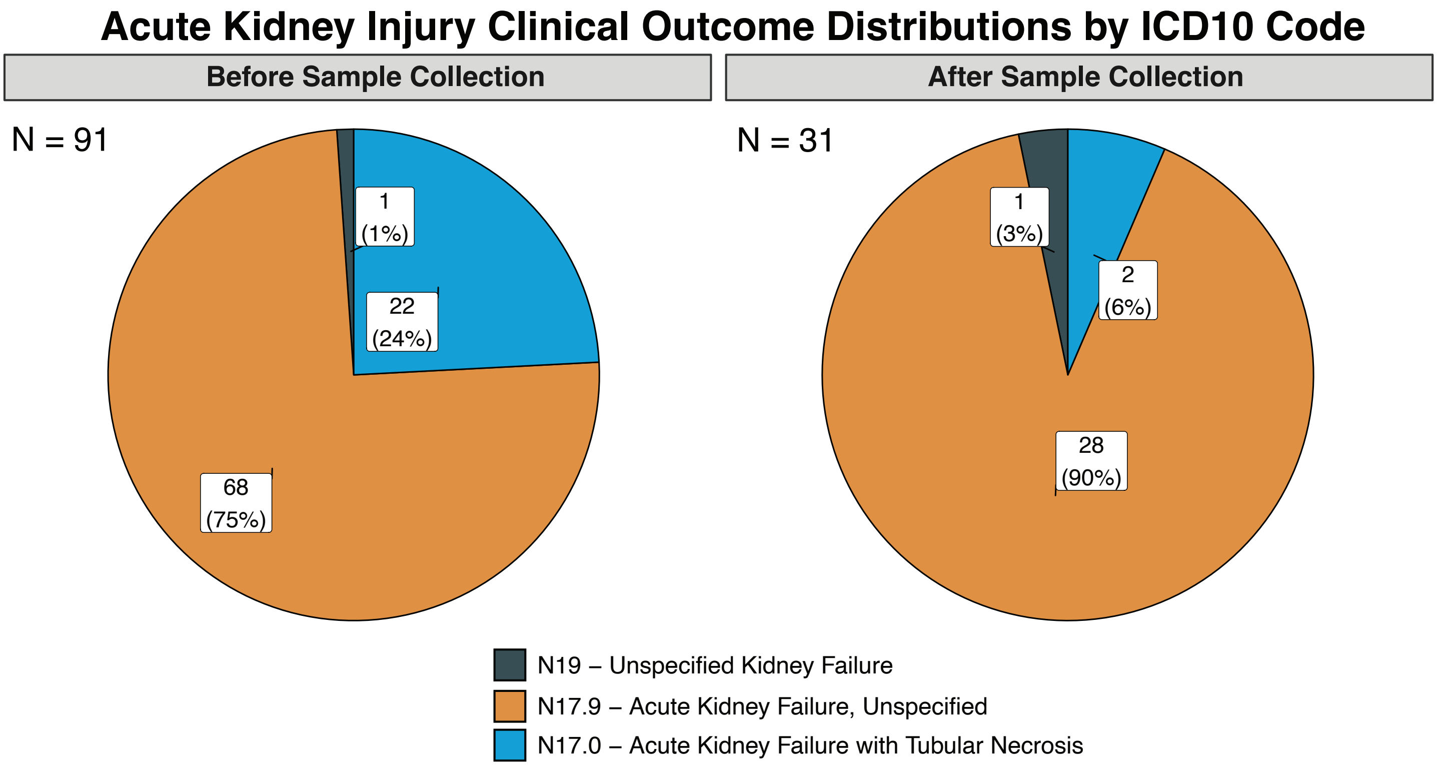


**Figure 4: Distribution of Acute Kidney Injury Clinical Outcomes**. ICD-10 code derived outcomes were assessed at two time-points: 1) the first acute kidney injury outcome found post COVID-19+ admission and prior to blood sample collection, 2) the first acute kidney injury outcome found post COVID-19+ admission, within 60-days after blood sample collection. Total outcome count for each time-point (N) are noted in the upper left hand corner of each pie-chart.

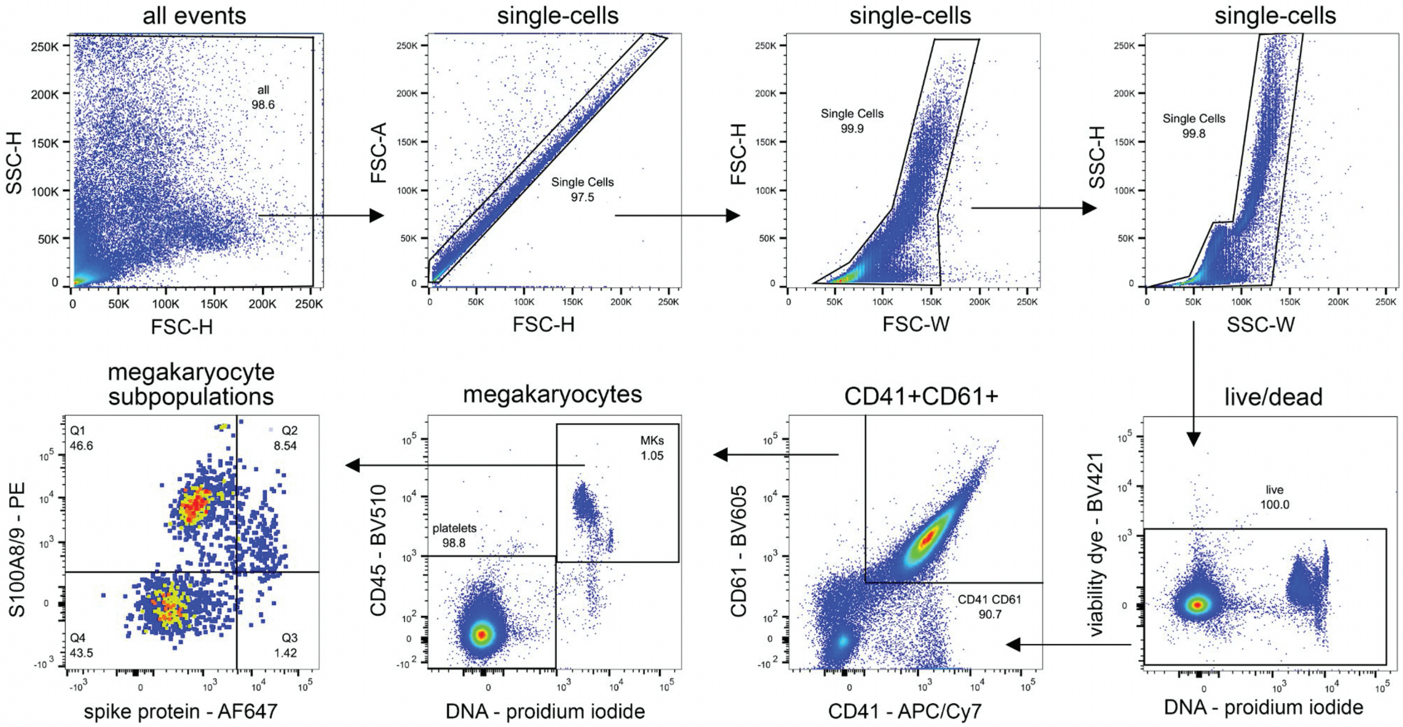


**Figure 5: Flow cytometry gating scheme for circulating megakaryocytes.**

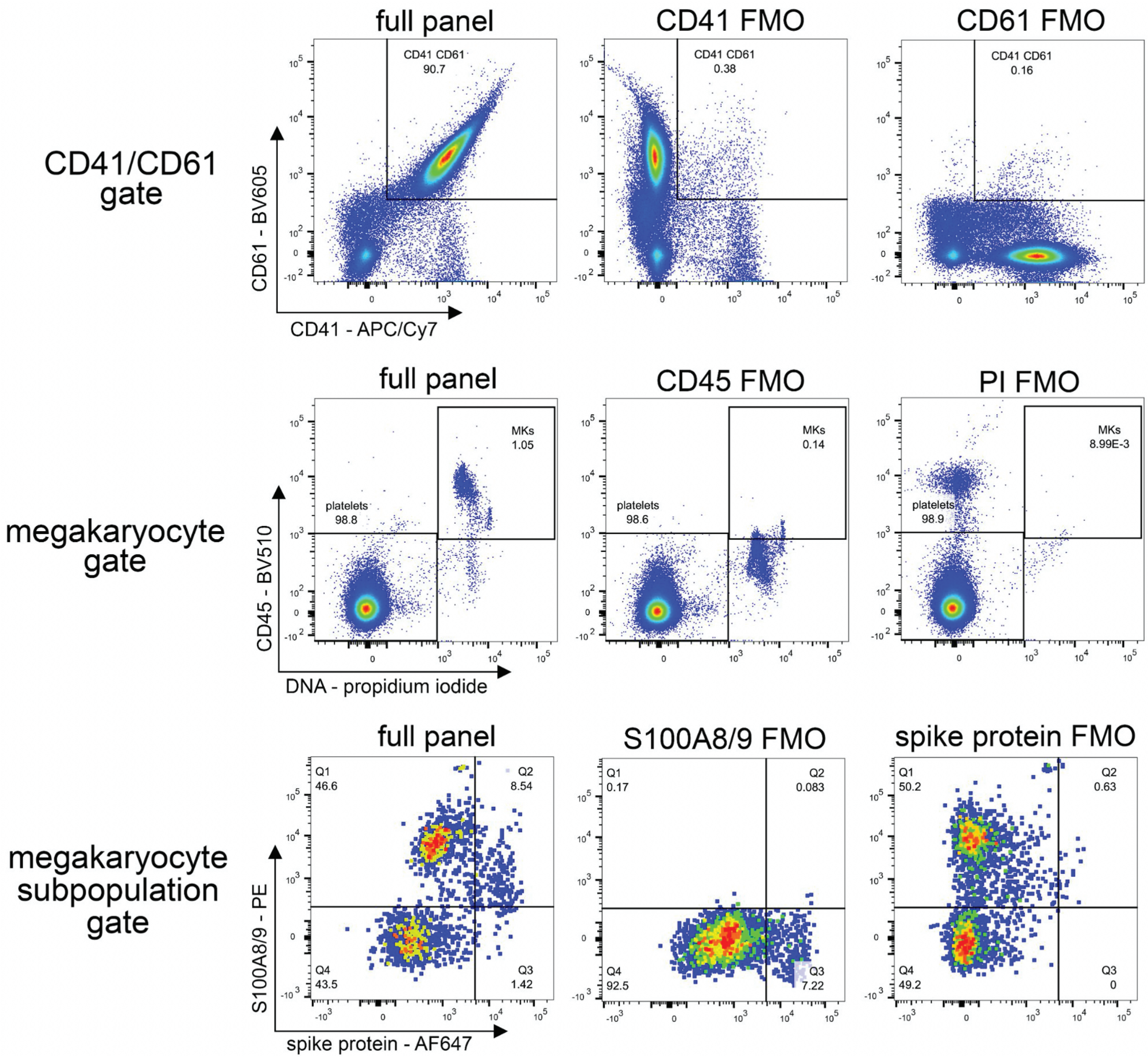


**Figure 6: Fluorescence minus one (FMO) flow cytometry controls.**

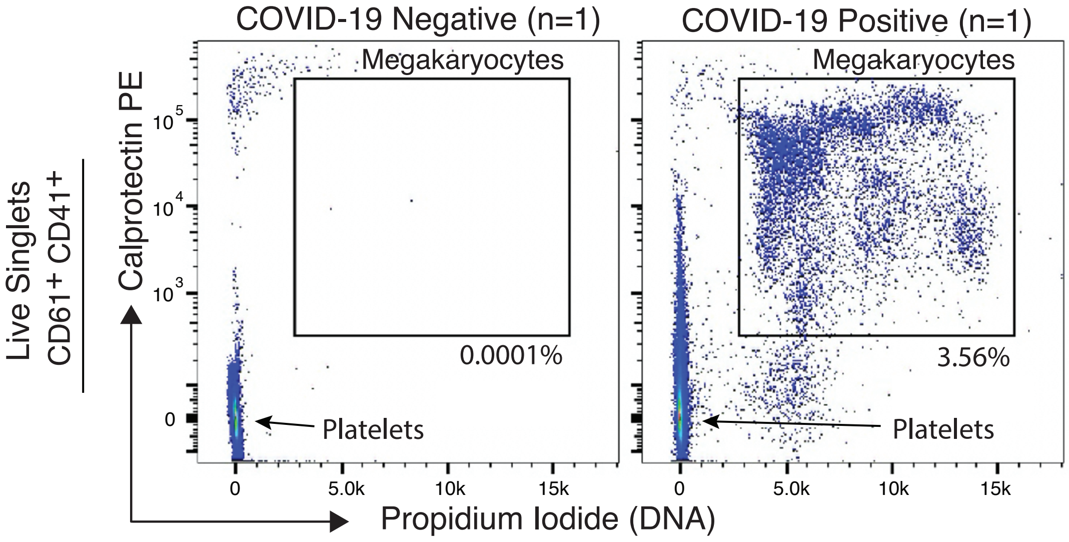


**Figure 7: Calprotectin expressing megakaryocytes in COVID-19 negative control vs. COVID-19 positive samples.** Flow cytometry plots of live singlets double-positive for CD61 and CD41 from COVID-19 negative control (n=1) and COVID-19 positive (n=1) peripheral blood UAB peripheral blood samples showing Calprotectin protein expression and propidium iodide (DNA) megakaryocytes in the plotting gates. DNA negative population in lower left corner of each plot are platelets.

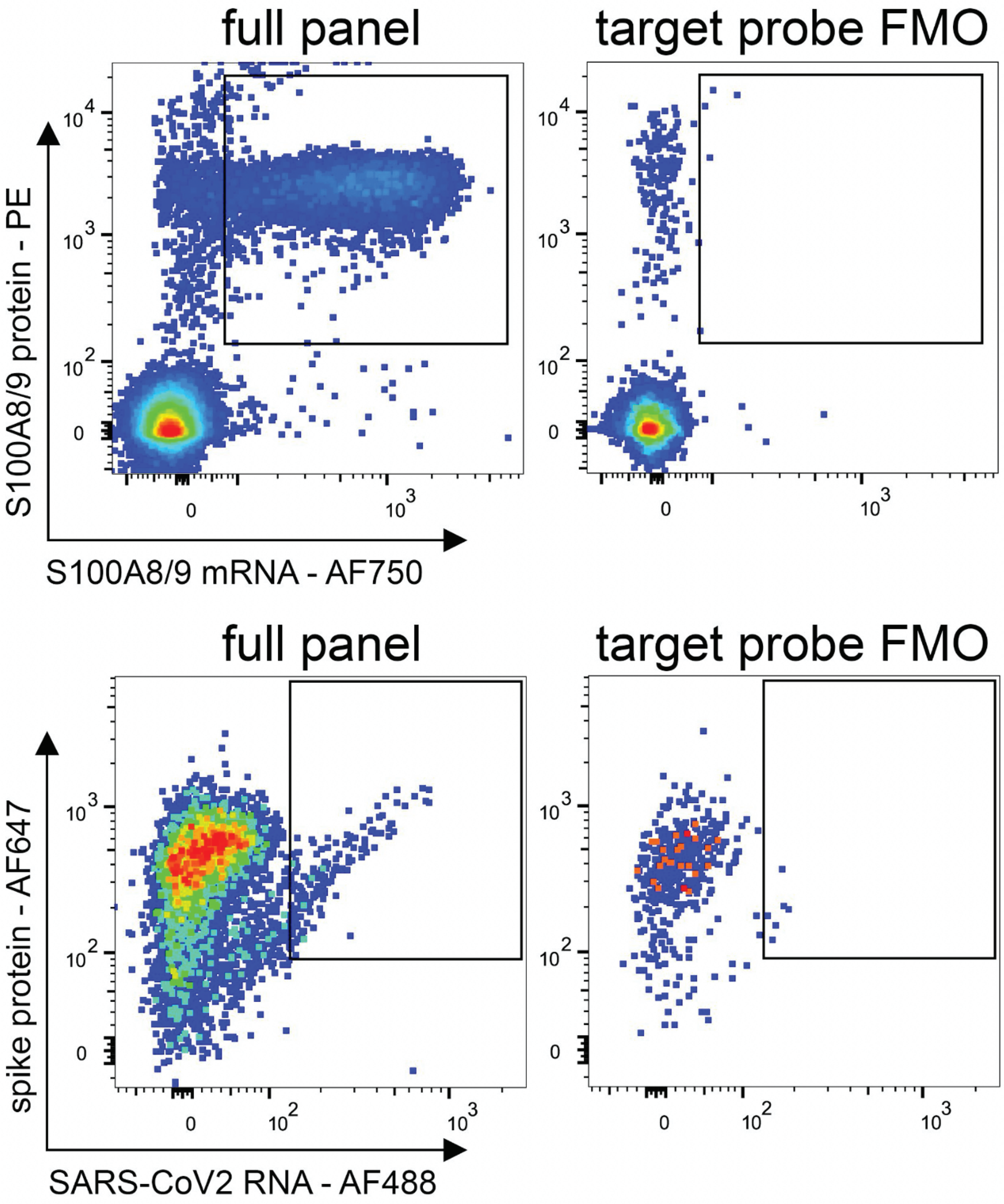


**Figure 8: Fluorescence minus one (FMO) PrimeFlow controls.** No target probe was used during the initial hybridization step for FMO controls. All other PrimeFlow steps were identical.

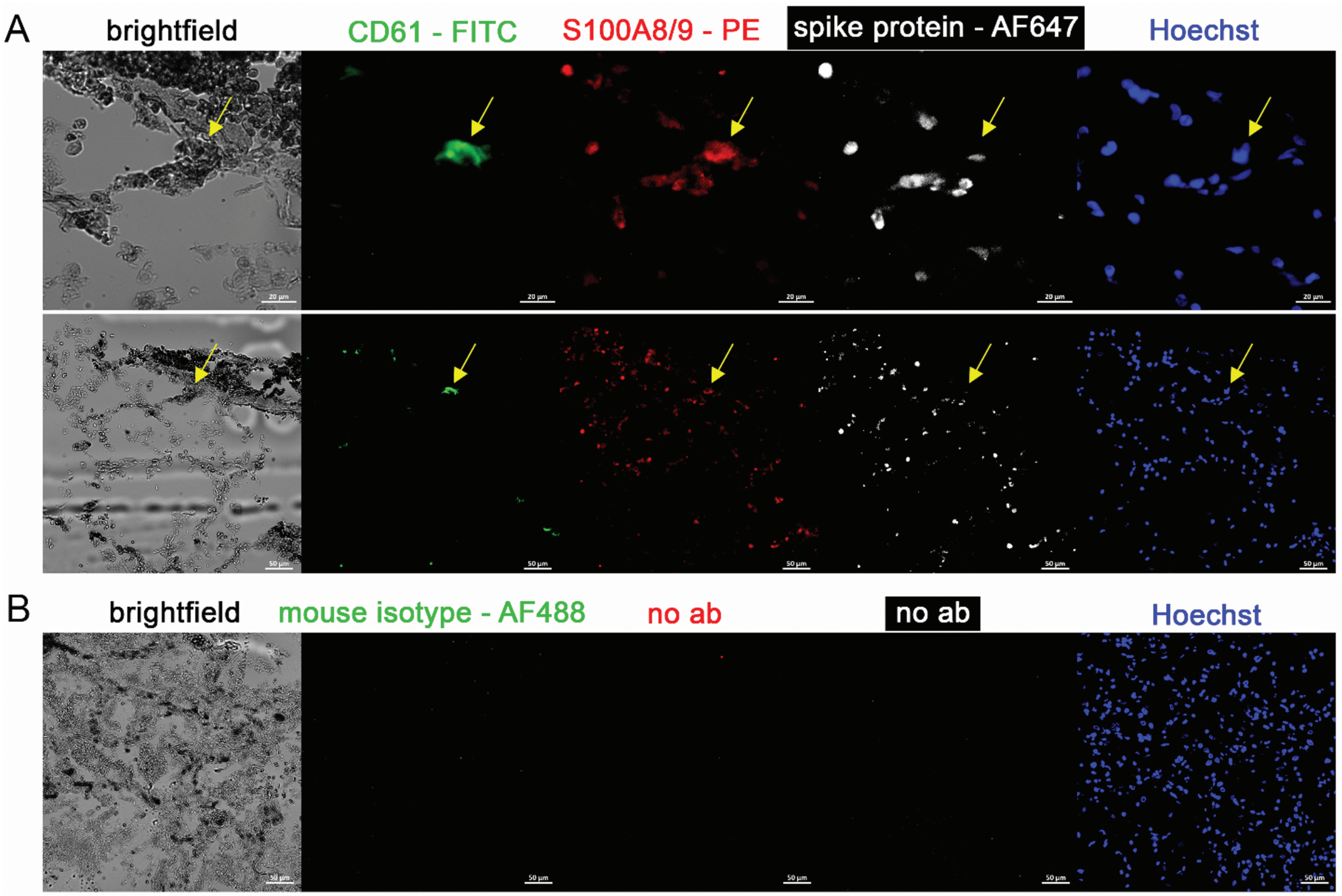


**Figure 9: Immunofluorescence of lung tissue from a COVID-19 patient with acute respiratory distress syndrome.** 5 channels are shown: brightfield (black pigment from TrueView autofluorescence quencher), green (CD61), red (S100A8/A9), white (spike protein), and blue (Hoechst). **(A)** Yellow arrow indicates megakaryocyte. Top row is 20X, bottom row is 60X. **(B)** Negative control stain using mouse isotype antibody.

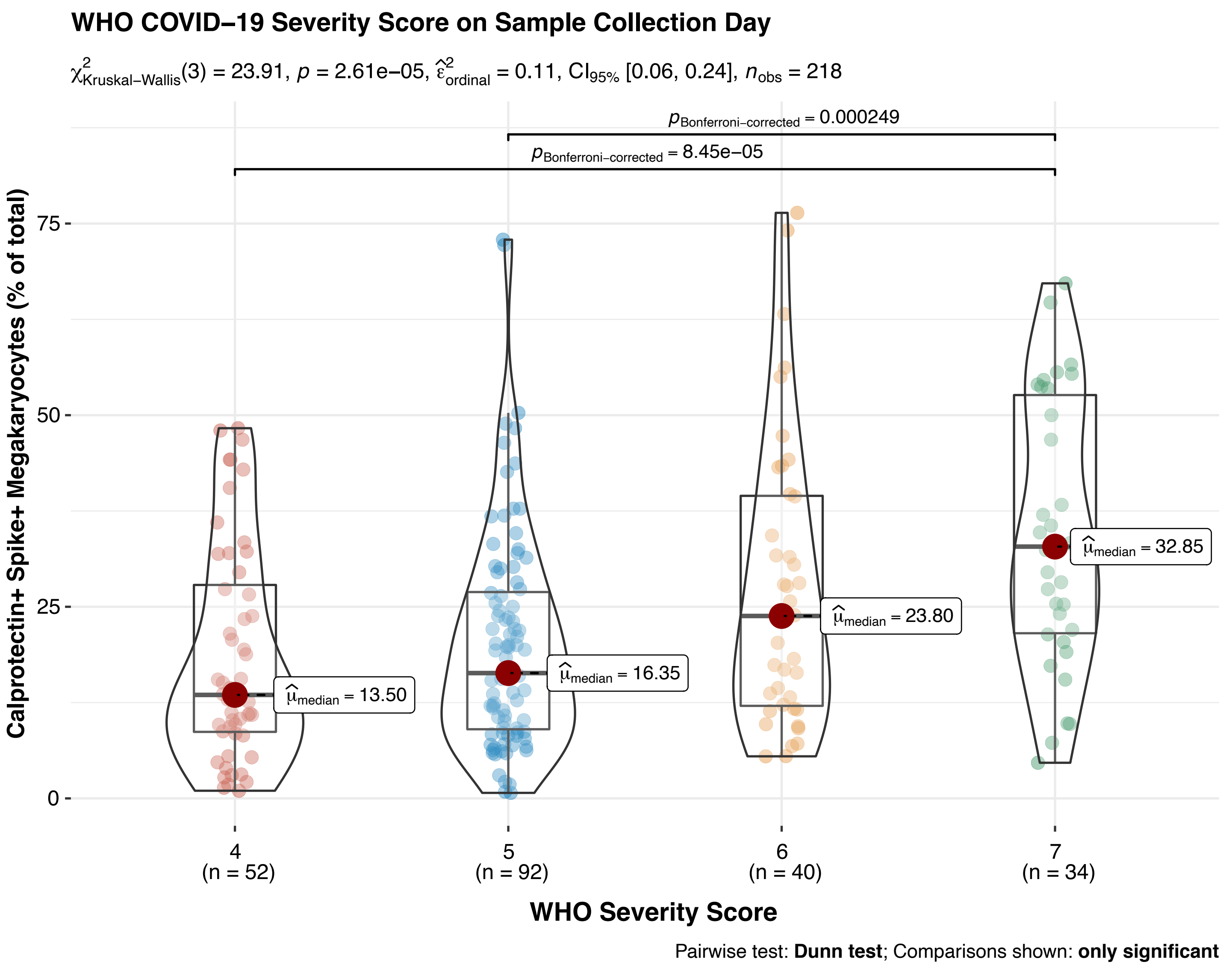


**Figure 10: Calprotectin+ Spike+ Megakaryocyte Proportion Positively Correlates with Peak WHO COVID-19 Severity Score on the Day of Sample Collection.**

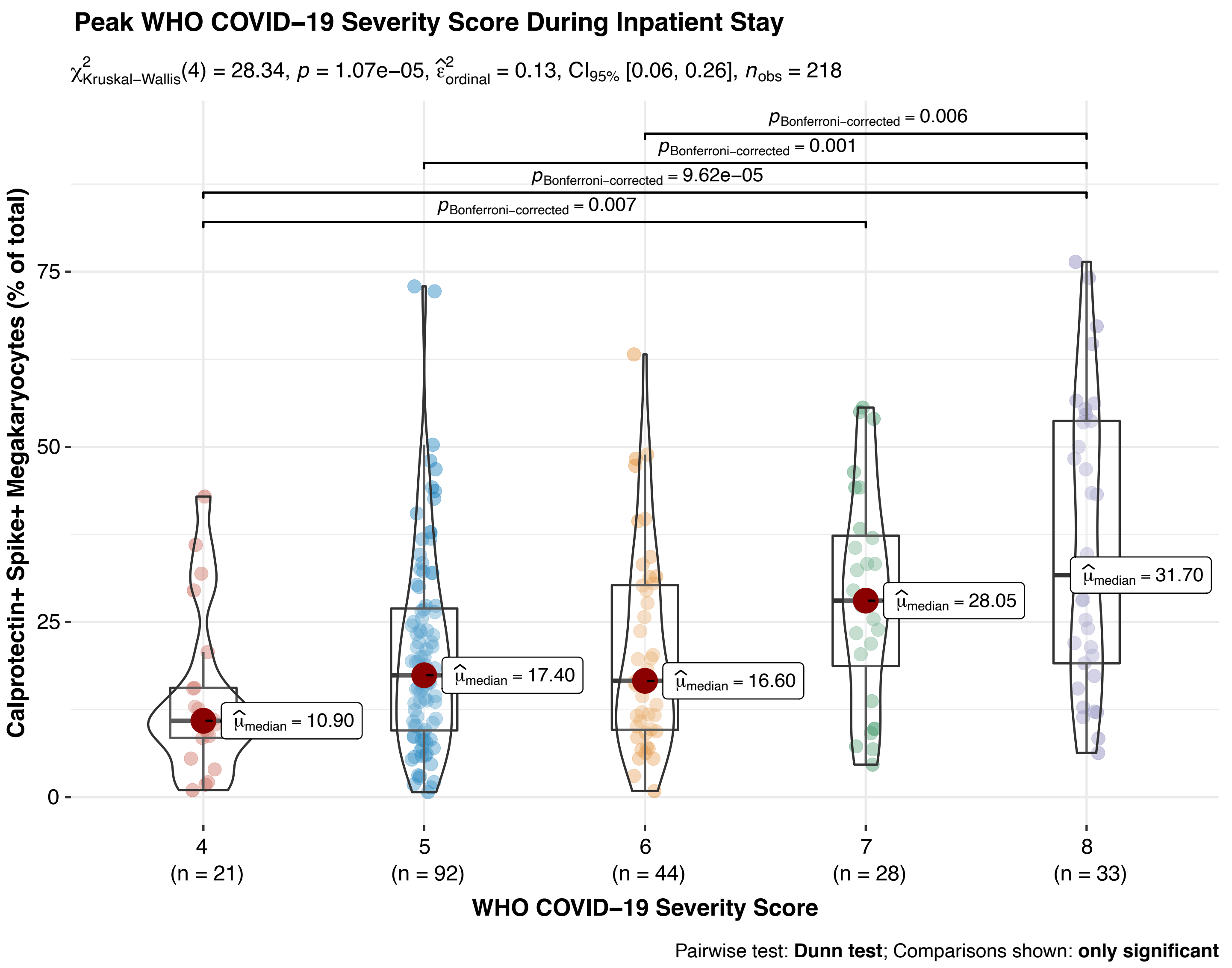


**Figure 11: Calprotectin+ Spike+ Megakaryocyte Proportion Positively Correlates with Peak WHO COVID-19 Severity Score During Inpatient Admission.**

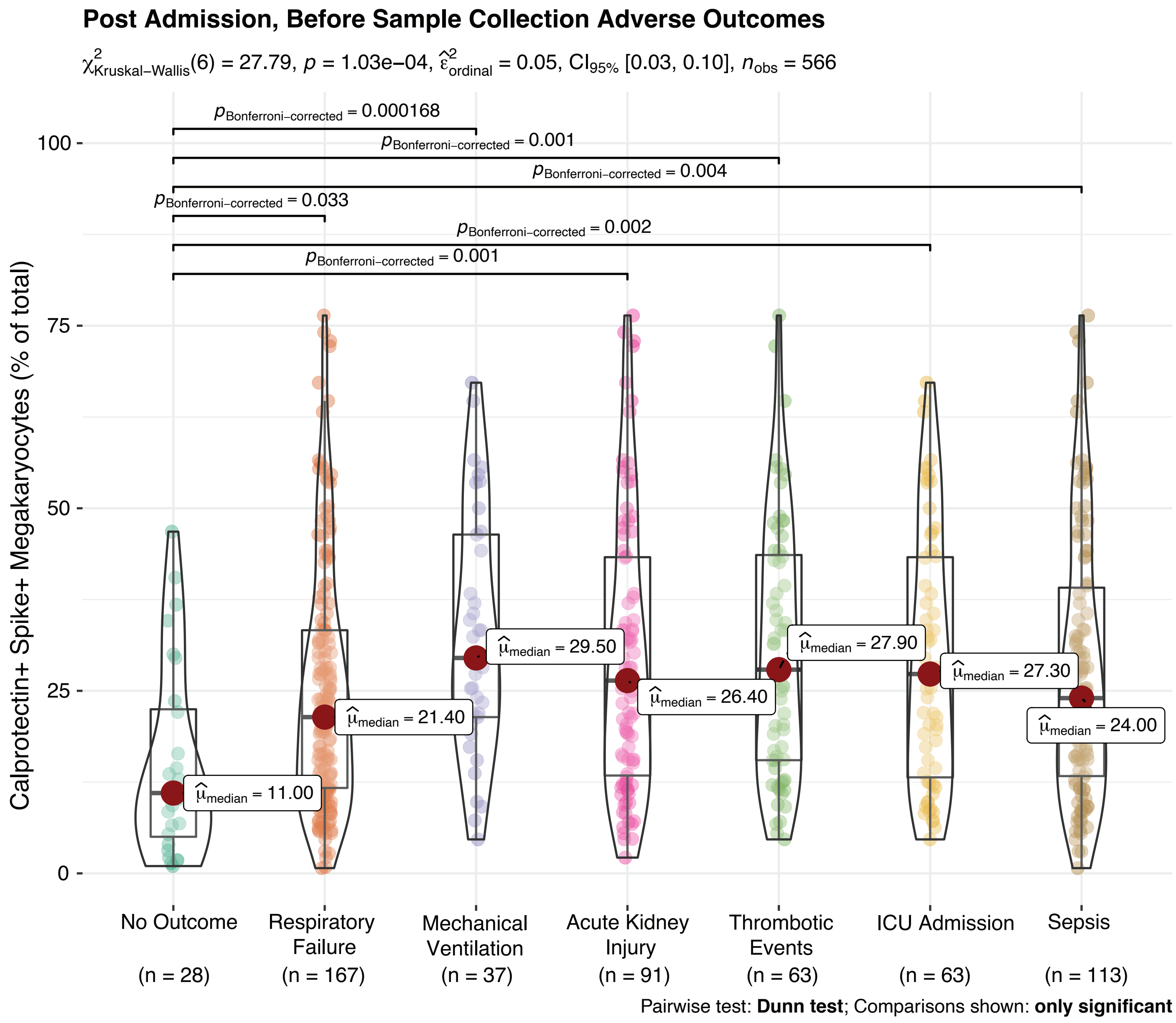


**Figure 12: Calprotectin+ Spike+ Megakaryocyte Proportion Positively Correlates with Adverse Outcomes Before Sample Collection.**

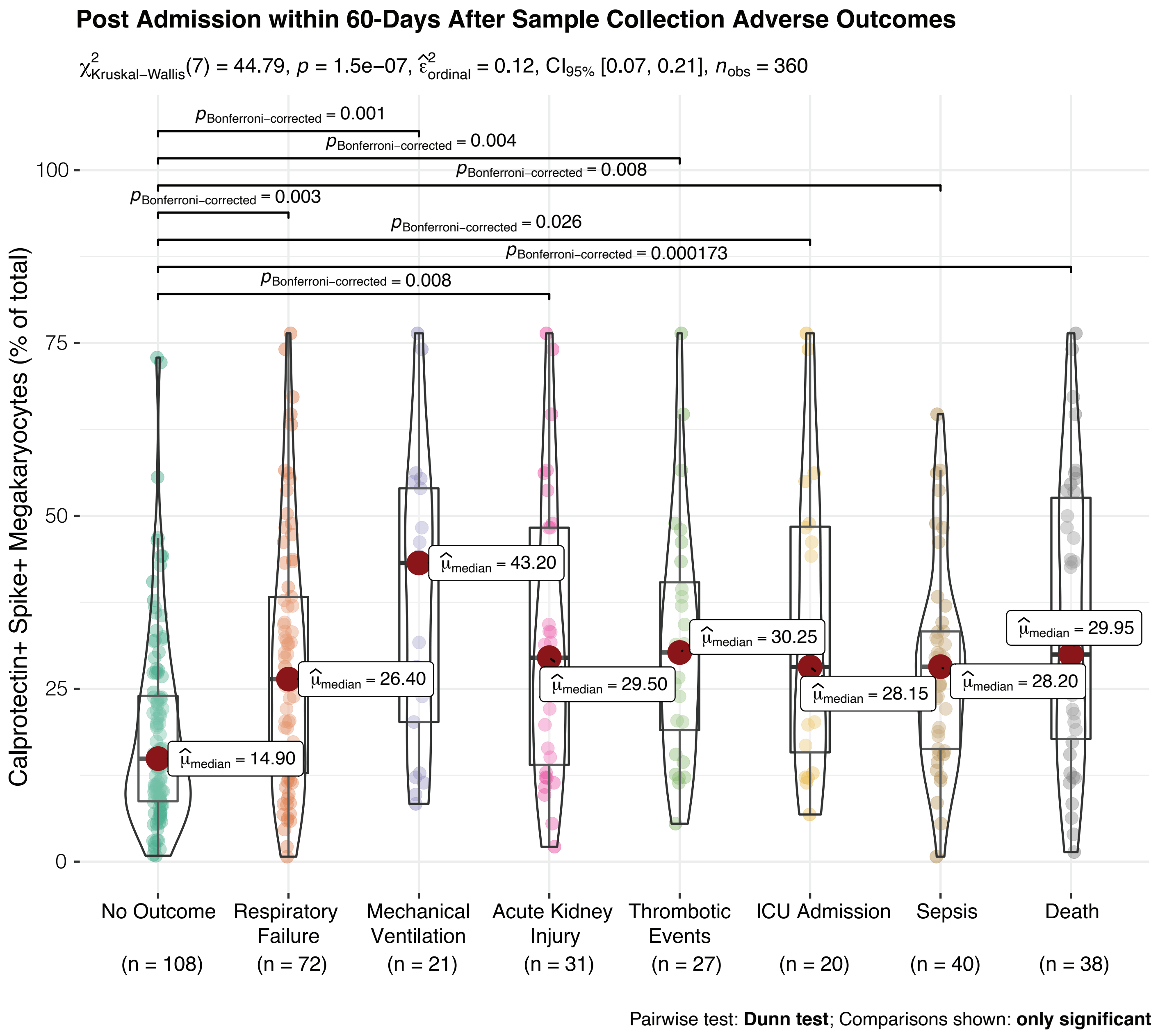


**Figure 13: Calprotectin+ Spike+ Megakaryocyte Proportion Positively Correlates with Adverse Outcomes 60-Days After Sample Collection.**

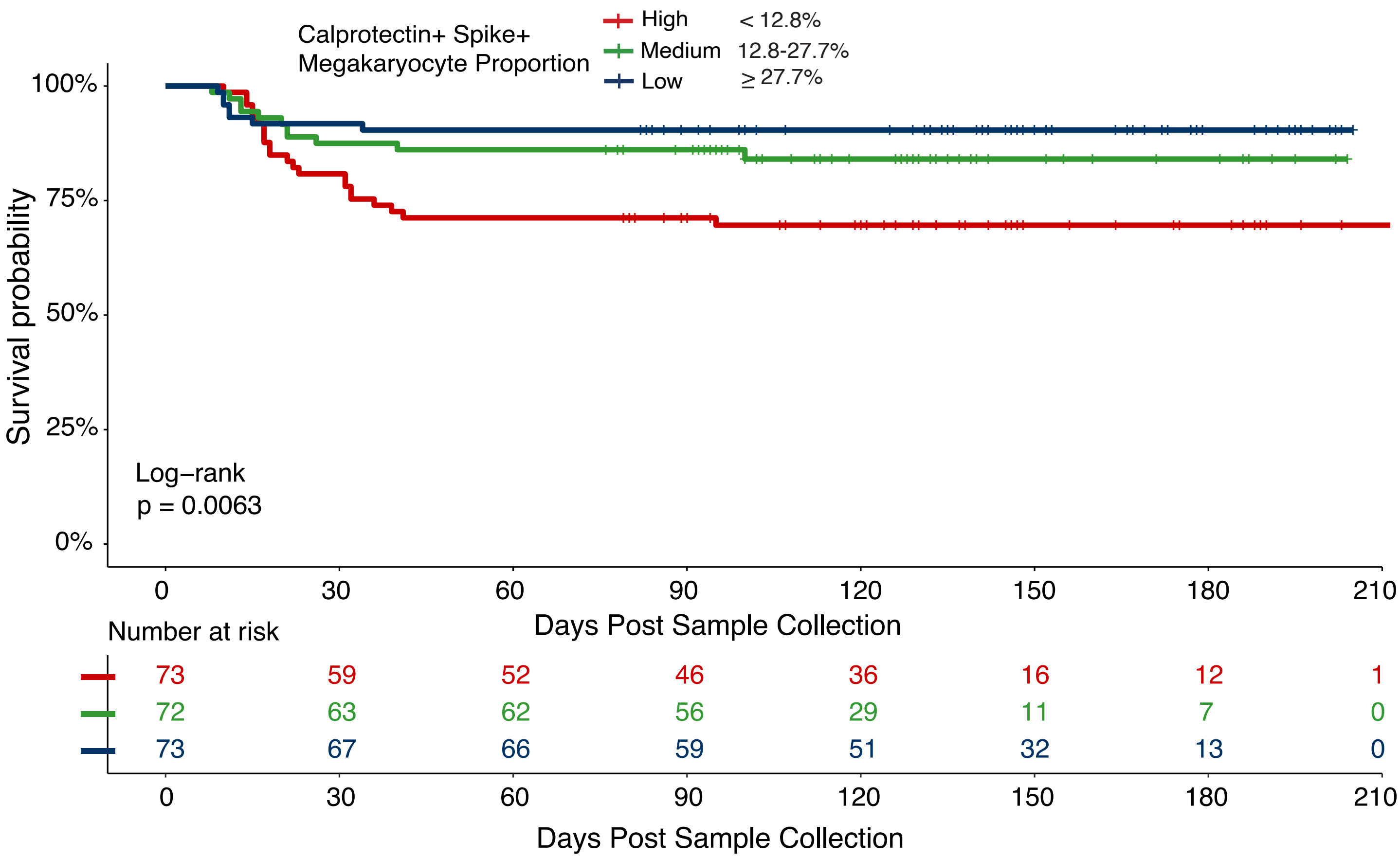


**Figure 14: Survival analysis stratified by calprotectin+ spike+ megakaryocyte proportion.**

| Variable | Adjusted Hazard Ratio (95% CI) | P value |
|---|---|---|
| Age | 1.02 (0.99-1.05) | 0.13 |
| Body Mass Index | 0.99 (0.95-1.04) | 0.74 |
| Time to Sample Collection | 1.01 (0.95-1.09) | 0.69 |
| Charlson Comorbidity Score | 1.10 (0.96-1.29) | 0.18 |
| Sex | | |
| *Male* | — | — |
| *Female* | 0.27 (0.11-0.66) | **0.004** |
| Calprotectin+ Spike+ Megakaryoctye Proportion | | |
| *Low (< 12.8%)* | — | — |
| *Medium (12.8-27.7%)* | 1.59 (0.50-4.13) | 0.34 |
| *High (≥ 27.7%)* | 2.50 (1.04-6.02) | **0.04** |

*For time to 60-day mortality, the hazard ratio was adjusted for age, sex, Charlson comorbidity score, and body mass index (BMI). A single patient from our cohort was ommitted from analysis due to a missing covariate (BMI) not recorded during their admission, results in n=217 patients for all models.

**Table 3: Multivariate Cox Regression for 60-Day Survival Stratified by calprotectin+ Spike+ Megakaryocyte Proportion.** Statistical significance for the Cox regression model was assessed using the companion applied regression analysis-of-variance (CAR-ANOVA) type III test.

| 30-Day Outcome | Event Rate | Unadjusted OR (95% CI) | P value | Adjusted OR (95% CI) | P value |
|---|---|---|---|---|---|
| All Cause Mortality | 13.36% | 2.23 (1.44-3.50) | <0.001 | 1.92 (1.23-3.05) | **0.004** |
| Mechanical Ventilation | 9.21% | 2.49 (1.51-4.18) | <0.001 | 2.41 (1.43-4.16) | **<0.001** |
| Respiratory Failure | 31.79% | 1.75 (1.24-2.50) | 0.001 | 1.64 (1.15-2.36) | **0.006** |
| Thrombotic Events | 11.52% | 1.78 (1.12-2.81) | 0.01 | 1.76 (1.09-2.85) | **0.02** |
| Acute Kidney Injury | 13.82% | 1.87 (1.21-2.88) | 0.004 | 1.70 (1.09-2.69) | **0.02** |
| ICU Admission | 18.43% | 1.44 (0.97-2.13) | 0.07 | 1.78 (1.03-3.06) | **0.03** |
| Sepsis | 8.75% | 1.93 (1.16-3.21) | 0.01 | 1.43 (0.94-2.15) | 0.08 |

*For all 30-day post sample collection outcomes, the odds ratio (OR) was adjusted for age, sex, Charlson comorbidity score and body mass index (BMI). Odds ratios represent increased likelihood of a given 30-day outcome for every 20% increase (20% shown) in Calprotectin+ Spike+ megakaryocyte proportion. A single patient from our cohort was ommitted from analysis due to a missing covariate (BMI) not recorded during their admission, results in n=217 patients for all models.

**Table 4: Multivariate Logistic Regression for 60-Day Survival Stratified by calprotectin+ Spike+ Megakaryocyte Proportion.** Statistical significance for the regression models were assessed using the companion applied regression analysis-of-variance (CAR-ANOVA) type III test.
